# Multilevel Factors Associated with Pediatric Leukemia: The Role of Rural Areas Adjacent to Urban Areas

**DOI:** 10.1101/2025.04.29.25326382

**Authors:** Benjamin Neil Vickers, April Rose Jimenez, Kristin Sarah Bogda

**Author notes:** **Corresponding Author** Benjamin Neil Vickers, Ph.D.

## Abstract

Leukemia is the most prevalent form of malignant cancer among children zero to 14 years of age. Established risk factors encompass a range of environmental exposures and patient attributes such as age, gender, and race/ethnicity; additionally, a correlation exists between lower socioeconomic status and an unfavorable prognosis. This research investigated whether attributes of residential counties in the United States consistently distinguished between pediatric leukemia patients and pediatric cancer patients without leukemia after accounting for various patient-level factors. The units of analysis consisted of pediatric (age < 20 years) cancer diagnoses reported to the Surveillance, Epidemiology, and End Results’ “SEER-21” cancer registry from 2010 through 2017 (N=44,808). The outcome was binary: cases were pediatric leukemia diagnoses (9.69%), and controls were the remaining non-leukemia pediatric cancer diagnoses. County-level predictors included urban-rural status, inflation-adjusted median household income, and proximity to tribal lands. Patient-level factors included age, sex, ethnicity/race, the reporting source, and the year of diagnosis. Using multilevel logistic regression, nonmetro counties adjacent to metro counties had 30 percent greater odds that a pediatric cancer was leukemia compared to counties of metro areas with over one million residents. The crude association between lower county median household income and pediatric leukemia was confounded by this nonmetro county adjacent to metro result. Males and all minorities also had higher odds that a given pediatric cancer would be diagnosed as leukemia. Exploration of these patient factors alongside documented environmental risk factors for pediatric leukemia should be conducted among residents of nonmetro counties neighboring metro counties. This urban-rural discrepancy in the odds of leukemia among pediatric cancer diagnoses should improve the identification of localized risks and prevention efforts in these communities.

## Introduction

Leukemia is the leading type of malignant pediatric cancer in the zero-to-14-year age range; acute lymphocytic leukemia (ALL) accounted for an estimated 26 percent, and acute myeloid leukemia (AML) accounted for an additional estimated 5 percent, of all new malignant pediatric cases in the U.S. in 2014 [1]. Additionally, a steady per year average increase of 0.7 percent has been recorded for the number of new leukemia cases in the U.S. in this age range since 1975 [2]. Although not as comparatively consequential in the 15-to-19-year age range, ALL and AML together accounted for an estimated 12 percent of new malignant cancers for this population of adolescents in the U.S. in 2014 [1].

Among the U.S. population from birth to under 20 years of age from 2006 through 2010, Hispanics had the highest age-adjusted leukemia incidence rate followed by non-Hispanic whites, non-Hispanic Asian/Pacific Islanders, and non-Hispanic blacks [1]. Even so, the mortality rate for pediatric non-Hispanic black leukemia patients was more similar to other groups due to a higher case-fatality rate [1]. Additionally, malignant pediatric cancers of all kinds have been shown to present significant and unforeseen financial and employment burdens on families and caregivers, especially for families in rural areas, and these burdens require special systems of support [3].

Pediatric leukemia in the U.S has been linked to genetics, the physical environment, and health behaviors in the current literature [4-6]. According to genetic research [7,8], around eight to ten percent or more of all children diagnosed with cancer had experienced a mutation in a predisposing gene that was inherited from the parents.

The observation of parental environmental exposures during prenatal and postnatal periods, as well as the child’s environmental exposures, has resulted in the identification of exposures to tobacco smoke, solvents, pesticides, other chemicals, ionizing radiation, and even traffic-caused air pollution as contributors to the development of pediatric leukemia [2]. Public health efforts aimed at reducing these exposures should likewise reduce the overall rate of childhood leukemia [2]. In addition to the significance of the type of exposure, whether it be environmental or genetic/molecular, the timing from exposure to pathogenesis is not necessarily immediate [9]. Notably, the fundamental molecular damage triggers for pediatric leukemia inherited in utero appear to delay their activation until infancy or childhood [9].

Since data have shown an increased risk of pediatric leukemia among Hispanics compared to non-Hispanic blacks and whites, this possibly implicates higher rates of harmful environmental exposures among Hispanic children and their parents [10]. In the large and ethnically diverse population of California, disproportionately high pediatric leukemia incidence has been observed for Hispanics, and this disparity has only been growing in recent years due to increases in Hispanic incidence rates [10].

While greater pediatric leukemia incidence rates have been observed among those of higher socioeconomic status [11], greater mortality rates have occurred in patients of lower socioeconomic status [12]. Inequitable access to healthcare services has had a noted impact on these disparities in pediatric leukemia outcomes [12]. Many lower socioeconomic status neighborhoods are simply not in close proximity to the necessary healthcare facilities [13]. Furthermore, treatments can be costly and are typically more accessible and affordable to those of higher socioeconomic status [14]. Similarly, while greater pediatric leukemia incidence rates have been observed in Hispanics and non-Hispanic whites, higher mortality rates have been observed for Hispanic and non-Hispanic black populations that tend to be more socially disadvantaged [1,15]. In short, vulnerable populations are disproportionally exposed to risk factors for cancer from early in life, so it is crucial that the disparities in exposures and their subsequent outcomes be addressed [16].

Given the current body of knowledge on this topic, there are two needs that the present study addressed. First, this study explored whether residential community-level factors were independently associated with whether pediatric cancer cases were diagnosed as leukemia after accounting for patient-level characteristics. Second, this study aimed to identify which urban-rural community categories in the U.S. might exhibit reliably greater odds that pediatric cancer cases were diagnosed as leukemia. The results of this study should advance our strategic public health knowledge and efforts on this topic toward identifying the types of communities where documented environmental and community-level risk factors appear to be contributing most consistently to pediatric leukemia incidence. Closing this gap is a necessary step toward confirming known, and identifying yet unknown, risk factors and high-risk populations for the implementation of corresponding community-based prevention strategies.

## Methods

This study had a cross-sectional case-control study design; the exposure/predictor data and case-control status were contemporaneous for each cancer diagnosis, which collectively made up the units of analysis for the study. The data were secondary; they were entrusted to the authors for research purposes free of charge by the National Cancer Institute’s (NCI) Surveillance, Epidemiology, and End Results (SEER) cancer registry. The study sample was a subset of the SEER 21 dataset limited to patient diagnosis years from 2010 through 2017, and patient age at diagnosis was limited to a maximum age of less than 20 years at the time of diagnosis. This resulted in a finite population sample of 44,808 pediatric cancer diagnoses observed over eight calendar years.

The SEER 21 dataset included cancer registries from 21 different reporting sources (see note*). The SEER 21 registry was exhaustive of all cancer diagnoses within the geographic coverage areas for each of the 21 reporting sources. The data were therefore representative of the populations of the combined mutually exclusive geographical areas that included some states and large metropolitan areas within the U.S. As a result, the sample for this study was not designed to represent the total pediatric population of the U.S., but nonetheless, the 21 registries covered sufficiently diverse parts of the U.S. to make the generalizability of the results of this study considerably strong for the U.S. pediatric population.

The study outcome was a binary pediatric cancer diagnosis variable. The variable was considered a case for a leukemia diagnosis (ICD-O-3 histology codes 9800-9949) and a control for any non-leukemia diagnosis. Eight variables were analyzed as potential predictors of a pediatric leukemia diagnosis. Patient-level demographic predictors included age in whole years, binary sex, and race/ethnicity in four categories: Hispanic of any race, non-Hispanic black, non-Hispanic white, and non-Hispanic of other or unknown race. Geographic/ecological predictors included eight ordinal inflation-adjusted (2018 U.S. Dollar) median household income categories of the county of residence (<$45k, $45k-<$50k, $50k-<$55k, $55k-<$60k, $60k-<$65k, $65k-<$70k, $70k-<$75k, $75k+), five ordinal categories of the rural/urban status of the county of residence based on the U.S. Department of Agriculture’s (USDA) [17] Rural-Urban Continuum Codes (RUCCs) (nonmetro county not adjacent to metro county, nonmetro county adjacent to metro county, county of metro area: population <250k, county of metro area: population 250k-1 million, county of metro area: >1 million), and binary purchased/referred care delivery area (PRCDA) (“1” for any county containing all or part of a tribal land/reservation or any county sharing a common boundary with a tribal land/reservation, “0” for all other counties). A nonmetro county is considered “adjacent” if it shares a boundary with a metro area county (or if a nonmetro noncore county shares a boundary with a micropolitan area county) and two percent or greater of the workforce commutes to the core of the adjacent metro area (or for a nonmetro noncore county, to the adjacent micropolitan area) [17]. Two other predictors were the calendar year of diagnosis (range: 2010-2017) and the binary reporting source (“1” for hospital inpatient/outpatient or physician’s office, “0” for other source, like a cancer care center).

Concerning any potential data bias, although the SEER-21 cancer registry dataset is large and has very strong external validity for the U.S. population, it does not collect data on all geographic areas of the U.S., so the results may be biased toward the geographies represented. Nonetheless, the percentages for the urban/rural-metro/nonmetro classifications in previous SEER data have matched acceptably well to these respective percentages for the total U.S. population [18]. The study design was cross-sectional, so no observed associations were considered to be causal. Even so, this study was conducted for the purpose of exploring possible causal hypotheses, so comments on how the results of this study corroborate prior etiological research, as well as discussions concerning possible etiologies suggested by these results, are included in the Discussion section. Last, the use of non-leukemia pediatric cancer controls means that this study examined differences among different types of pediatric cancers. Therefore, this study does not contribute to the comparison of pediatric leukemia cases to the larger pediatric population that was cancer-free. Nonetheless, differences observed between pediatric leukemia and non-leukemia cancer cases may have important implications for possible differential etiologies between these groups.

Each of the eight predictors was described according to the appropriate measure of central tendency and distribution for pediatric leukemia cases as a group and for the non-leukemia pediatric cancer controls as a group, and the appropriate statistical test of bivariate associations were performed using SAS Version 9.4 (https://www.sas.com). These included the chi-square test for nominal predictors and the Cochran-Armitage test for the single ordinal predictor: inflation-adjusted median household income of the county of residence (Table 1).

**Table 1.** Descriptive Statistics: pediatric (under 20 years) leukemia cases and non-leukemia cancer controls by patient-level and community-level characteristics (SEER 21, 2010-2017)

| Predictors | Leukemia Cases<br>n=4,342 (9.69%) | Non-Leukemia Controls<br>n=40,466 (90.31%) | p-value |
| --- | --- | --- | --- |
| *{$\chi^2$ test & Cochran-Armitage trend Z-test: count (column %)}* |  |  |  |
| Age Category (none missing) $\{\chi^2 = 79.7313\}$ | | | <0.001 |
| <1 year | 311 (7.16%) | 2,484 (6.14%) |  |
| 1 – 4 years | 974 (22.43%) | 9,375 (23.17%) |  |
| 5 – 9 years | 821 (18.91%) | 6,901 (17.05%) |  |
| 10 – 14 years | 1,018 (23.45%) | 8,026 (19.83%) |  |
| 15 – 19 years | 1,218 (28.05%) | 13,680 (33.81%) |  |
| Sex (none missing) $\{\chi^2 = 75.54\}$ | | | <0.001 |
| Male | 2,567 (59.12%) | 21,120 (52.19%) |  |
| Female | 1,775 (40.88%) | 19,346 (47.81%) |  |
| Ethnicity/Race (none missing) $\{\chi^2 = 67.36\}$ | | | <0.001 |
| Hispanic (any race) | 1,256 (28.93%) | 11,207 (27.69%) |  |
| Non-Hispanic (NH) Black | 567 (13.06%) | 4,189 (10.35%) |  |
| Non-Hispanic (NH) White | 2,024 (46.61%) | 21,189 (52.36%) |  |
| NH (other/unknown race) | 495 (11.40%) | 3,881 (9.59%) |  |
| Reporting Source (none missing) $\{\chi^2 = 23.52\}$ | | | <0.001 |
| Hospital or Clinic | 4,196 (96.64%) | 38,431 (94.97%) |  |
| Other (e.g., cancer center) | 146 (3.36%) | 2,035 (5.03%) |  |
| Year of Diagnosis (none missing) $\{\chi^2 = 16.36\}$ | | | 0.022 |
| 2010 | 539 (12.41%) | 4,951 (12.23%) |  |
| 2011 | 504 (11.61%) | 5,079 (12.55%) |  |
| 2012 | 520 (11.98%) | 5,054 (12.49%) |  |
| 2013 | 568 (13.08%) | 4,972 (12.29%) |  |
| 2014 | 565 (13.01%) | 5,109 (12.63%) |  |
| 2015 | 519 (11.95%) | 5,308 (13.12%) |  |
| 2016 | 544 (12.53%) | 5,114 (12.64%) |  |
| 2017 | 583 (13.43%) | 4,879 (12.06%) |  |
| Urban-Rural County Status (missing = 0.21%) $\{\chi^2 = 11.38\}$ | | | 0.023 |
| Nonmetro not next to metro | 144 (3.33%) | 1,339 (3.32%) |  |
| Nonmetro next to metro | 266 (6.15%) | 2,025 (5.01%) |  |
| Metro: < 250,000 | 291 (6.72%) | 2,594 (6.42%) |  |
| Metro: 250,000-1 million | 858 (19.82%) | 8,067 (19.97%) |  |
| Metro: > 1 million | 2,769 (63.98%) | 26,363 (65.27%) |  |
| County Contains or Next to Tribal Land [PRCDA] (missing = 0.02%) $\{\chi^2 = 4.00\}$ | | | 0.046 |
| Yes | 1,282 (29.54%) | 12,549 (31.02%) |  |
| No | 3,058 (70.46%) | 27,912 (68.98%) |  |
| County Median Household Income (inflation-adjusted) (missing = 0.02%) $\{Z = 3.3896\}$ | | | <0.001 |
| < \$45,000 | 431 (9.93%) | 3,678 (9.09%) | |
| \$45,000-\$49,999 | 304 (7.00%) | 2,552 (6.31%) | |
| \$50,000-\$54,999 | 436 (10.05%) | 4,039 (9.98%) | |
| \$55,000-\$59,999 | 535 (12.33%) | 4,985 (12.32%) | |
| \$60,000-\$64,999 | 812 (18.71%) | 7,167 (17.71%) | |
| \$65,000-\$69,999 | 376 (8.66%) | 3,736 (9.23%) | |
| \$70,000-\$74,999 | 291 (6.71%) | 2,555 (6.31%) | |
| ≥ \$75,000 | 1,155 (26.61%) | 11,749 (29.04%) | |

Note: Significant *p*-values using Bonferroni adjustment ( $\alpha_j=0.05/8=0.00625$ ) appear in *italics*.

Multilevel logistic regression modelling (SAS 9.4; https://www.sas.com) was used to observe the adjusted odds ratios for the eight predictor variables. All predictor variables were retained in the model (Table 2) due to the retention of each one resulting in an improved model fit (i.e., lower Akaike Information Criterion). There were 22 regression model parameters across eight separate predictors. Using the standard statistical power of 0.8 and the Type 1 error probability (α) of 0.05, the model required a minimum sample size of 2,170 to detect a multiple correlation coefficient of 0.1 [19,20]. Given that all eight predictors were tested equally for their associations with the outcome in the multilevel model, the Bonferroni adjustment to the Type 1 error probability was made to account for eight null hypotheses, one for each predictor (α_j_=0.05/8=0.00625), which preserved the model Type 1 error probability of 0.05 [21].

**Table 2.**
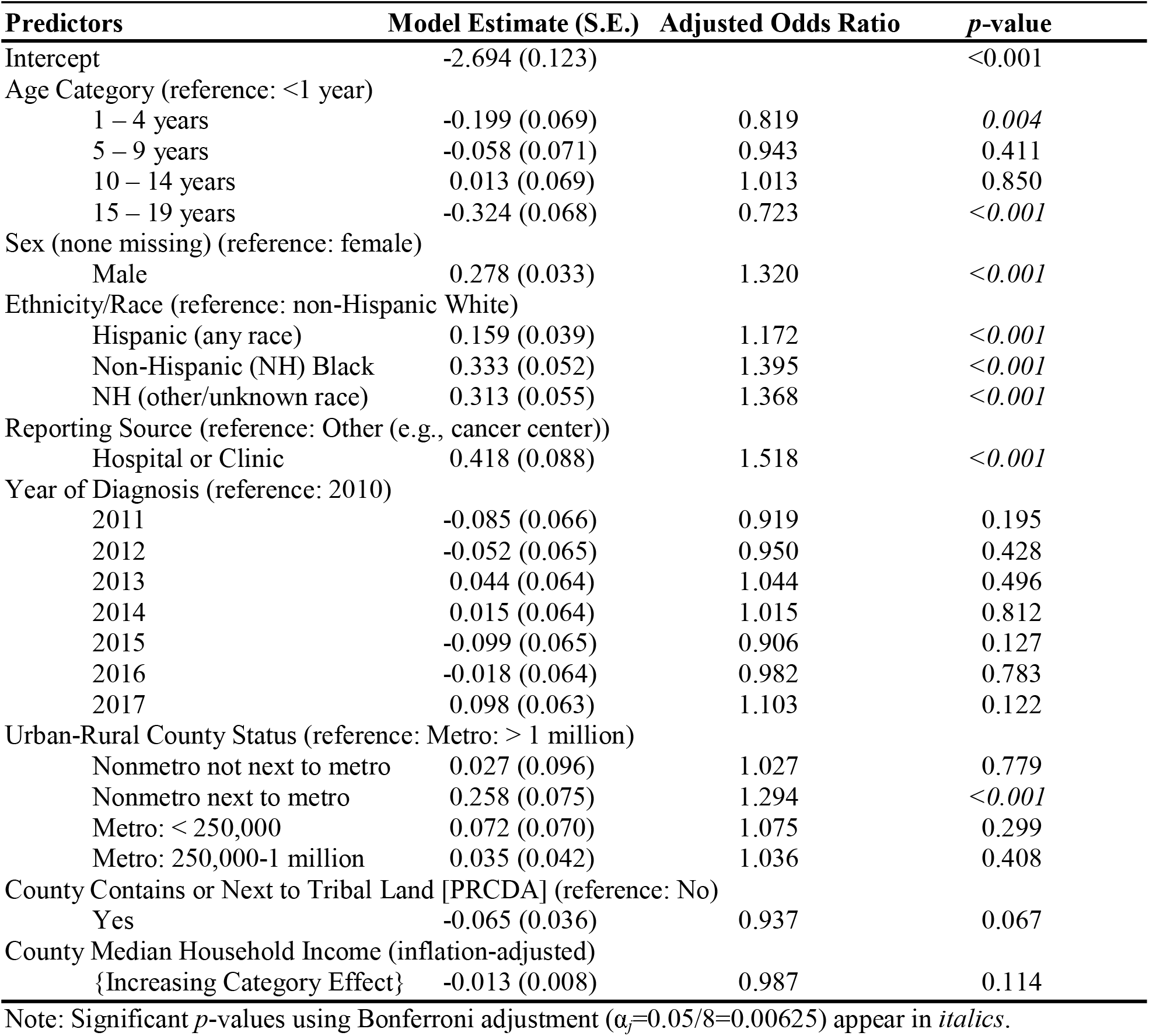
Multilevel Logistic Regression Model: adjusted odds ratios for patient-level and community-level predictor associations with pediatric (under 20 years) leukemia cases versus pediatric non-leukemia cancer controls (SEER 21, 2010-2017)

| Predictors | Model Estimate (S.E.) | Adjusted Odds Ratio | p-value |
| --- | --- | --- | --- |
| Intercept | -2.694 (0.123) |  | <0.001 |
| Age Category (reference: <1 year) |  |  |  |
| 1 – 4 years | -0.199 (0.069) | 0.819 | *0.004* |
| 5 – 9 years | -0.058 (0.071) | 0.943 | 0.411 |
| 10 – 14 years | 0.013 (0.069) | 1.013 | 0.850 |
| 15 – 19 years | -0.324 (0.068) | 0.723 | <0.001 |
| Sex (none missing) (reference: female) |  |  |  |
| Male | 0.278 (0.033) | 1.320 | <0.001 |
| Ethnicity/Race (reference: non-Hispanic White) |  |  |  |
| Hispanic (any race) | 0.159 (0.039) | 1.172 | <0.001 |
| Non-Hispanic (NH) Black | 0.333 (0.052) | 1.395 | <0.001 |
| NH (other/unknown race) | 0.313 (0.055) | 1.368 | <0.001 |
| Reporting Source (reference: Other (e.g., cancer center)) |  |  |  |
| Hospital or Clinic | 0.418 (0.088) | 1.518 | <0.001 |
| Year of Diagnosis (reference: 2010) |  |  |  |
| 2011 | -0.085 (0.066) | 0.919 | 0.195 |
| 2012 | -0.052 (0.065) | 0.950 | 0.428 |
| 2013 | 0.044 (0.064) | 1.044 | 0.496 |
| 2014 | 0.015 (0.064) | 1.015 | 0.812 |
| 2015 | -0.099 (0.065) | 0.906 | 0.127 |
| 2016 | -0.018 (0.064) | 0.982 | 0.783 |
| 2017 | 0.098 (0.063) | 1.103 | 0.122 |
| Urban-Rural County Status (reference: Metro: > 1 million) |  |  |  |
| Nonmetro not next to metro | 0.027 (0.096) | 1.027 | 0.779 |
| Nonmetro next to metro | 0.258 (0.075) | 1.294 | <0.001 |
| Metro: < 250,000 | 0.072 (0.070) | 1.075 | 0.299 |
| Metro: 250,000-1 million | 0.035 (0.042) | 1.036 | 0.408 |
| County Contains or Next to Tribal Land [PRCDA] (reference: No) |  |  |  |
| Yes | -0.065 (0.036) | 0.937 | 0.067 |
| County Median Household Income (inflation-adjusted) |  |  |  |
| {Increasing Category Effect} | -0.013 (0.008) | 0.987 | 0.114 |

Note: Significant *p*-values using Bonferroni adjustment ( $\alpha=0.05/8=0.00625$ ) appear in *italics*.

The greatest percentage of missing values among the selected variables in the final analytic sample was for the urban/rural status variable (0.21% missing). Given the negligible missing percentages for urban/rural status and the other variables, patients with missing values were excluded from the analysis. Furthermore, given the external validity and robust data collection from the many comprehensive cancer registries across several urban area and statewide jurisdictions, case-control matching and sensitivity analyses were not used in the collection and analysis of the data.

### Ethics Approval

The SEER-21 cancer registry data were provided (see note†) for this study as a de-identified and limited secondary dataset. Since these data did not qualify as human subjects research data in this form, our use of the data did not require an ethics review by an Institutional Review Board.

## Results

The November 2019 version of SEER 21 includes 9,821,960 records across 18 years (2000-2017). According to the National Cancer Institute [22], approximately one percent of all cancer diagnoses occur in the under 20 years of age category. Restricting the data to an age of less than 20 years at the time of diagnosis, the data were reduced to 97,998 records, which is one percent (0.998%) of the records in the full dataset. There were significant changes to leukemia diagnostic coding for cases diagnosed from January 1, 2010 forward [23], so the dataset was restricted to cases diagnosed from 2010 through 2017. This decision reduced the analytic sample to a final count of 44,808 pediatric cancer cases. Of these pediatric cancer diagnoses, almost one in ten (4,342 cases, 9.69%) had a leukemia code. This large finite population sample was more than minimally powered for the multilevel logistic modelling discussed in the Methods. Additionally, since non-leukemia controls far outnumbered leukemia cases, which were much closer to the required minimum sample size, large sample p-value scaling was not used. Furthermore, the use of the Bonferroni correction largely removes the need for large sample scaling [21].

Table 1 shows the descriptive statistics for the leukemia cases and non-leukemia controls, and the results of statistical tests of bivariate associations of cases and controls with the predictors are shown. Using the Bonferroni adjustment, five of the eight predictors had significant unadjusted associations with pediatric leukemia diagnoses. Compared to controls, pediatric leukemia diagnoses occurred more among infants and the five-to-fourteen-year age range, were reported more from hospitals and clinics, and were more common among males and racial/ethnic minorities. Among ecological factors, pediatric leukemia made up greater proportions of pediatric cancer cases in counties with lower median household incomes.

In the multilevel logistic regression model (Table 2), the age category, sex, ethnicity/race, reporting source, and urban-rural county status were significantly associated with pediatric leukemia diagnoses. The adjusted odds of a pediatric cancer diagnosis being leukemia were significantly smaller in the one-to-four-year and 15-to-19-year age ranges than were observed among infants. The adjusted odds for the five-to-14-year age range were not significantly different from those of infants. Furthermore, the odds of the patient being male were 32 percent higher among leukemia diagnoses compared to other pediatric cancers. Likewise, the odds of any minority race/ethnicity were significantly higher among leukemia diagnoses, especially for a non-Hispanic ethnicity with black, other, or unknown race. The largest model effect was observed for the reporting source: pediatric leukemia diagnoses had odds of being reported by a hospital or clinic, as opposed to other reporting sources, that were 52 percent greater than those observed for the other pediatric cancer diagnoses. Last, while no ordinal effect was observed for county urban-rural classification, the odds of a pediatric leukemia diagnosis were almost 30 percent higher in nonmetro counties that were adjacent to metro counties compared to counties of metro areas with over 1 million residents. The odds of pediatric leukemia were undifferentiated from the odds of other pediatric cancers among all other urban-rural classifications compared to metro areas with over 1 million residents.

A lower county median household income, which was associated with higher percentages of pediatric cancer diagnoses that were leukemia in the crude bivariate analysis (Table 1), became a non-significant predictor in the multilevel model due to confounding with urban-rural county status. This confounding was driven by nonmetro counties tending to have lower median household incomes. No meaningful or directional trend or period effects were observed over the timeframe of the data collection. To reiterate from the Methods, the retention of all eight predictors in the multilevel logistic regression improved model fit.

## Discussion

This study found that social determinants at both the patient level and the aggregated county level were independently associated with the likelihood that a pediatric cancer was diagnosed as leukemia. Due to leukemia being a cancer of the blood, it was not surprising that the odds were greater for pediatric leukemia diagnoses being reported by a regular hospital or clinic, where blood tests are readily available, than was observed for other pediatric cancers. Among patient-level factors, the adjusted odds that a pediatric cancer was diagnosed as leukemia were relatively higher for infants, five-to-14-year-olds, males, and all minority race/ethnicity categories compared to non-Hispanic white patients. In county-level factors, nonmetro counties that were adjacent to metro counties were associated with significantly greater odds of a leukemia diagnosis when compared to other county types. The unadjusted association of lower county median household income with pediatric leukemia was confounded with this stronger effect observed for nonmetro counties adjacent to metro counties, as these counties tended to have lower median household incomes than metro counties. Furthermore, these generalizable results exhibited a robust pattern over the 2010 to 2017 pooled data timeframe, as the marginal odds of a leukemia diagnosis associated with the calendar year did not significantly deviate in any of the years from 2011 to 2017 from those in 2010, the baseline collection year.

An important contribution of this study was the examination of contextual community factors alongside patient-specific characteristics simultaneously in a multilevel-adjusted analysis. Additionally, this analysis examined the role of rural-urban communities in pediatric leukemia diagnosis odds based on the USDA’s [17] RUCCs classification available in the analytic SEER 21 dataset. Compiling all these factors as predictors within a single study intentionally followed the prescribed methodology of Meilleur et al. [24], making this study a prescribed methodological advancement in the area of urban-rural cancer research.

Using the RUCCs classification system, Delavar et al. [25] found no significant differences in survival hazard ratios based on metro versus nonmetro nor urban versus rural areas of residence among pediatric cancer patients. However, that study did not examine the specific cancer type, which might have been consequential, as it was in the present study. For example, Blake et al. [18] found higher incidence and mortality rates among “rural” (i.e., nonmetro, based on RUCCs) populations for certain types of cancers such as “cervical cancer (measured among women only) as well as colorectal, kidney, lung and bronchus, melanoma, and oropharyngeal cancers.” On the other hand, Blake et al. [18] found higher incidence rates among metro populations for liver, thyroid, and breast cancers. While the present study did not examine incidence nor mortality rates, significantly greater odds that a pediatric cancer was diagnosed as leukemia were observed for nonmetro counties, but only among those that were adjacent to metro areas. The almost 30 percent greater odds are noteworthy given that these are marginal model odds after accounting for significant patient-level characteristics like age, sex, race/ethnicity, and reporting source. Similarly, among nonmetro areas with urbanized (based on population density) populations of 2,500 or more, Blake et al. [18] observed higher cancer incidence rates for those counties that were adjacent to metro areas. Combining the Blake et al. study [18] with the present study, the adjacency of nonmetro communities to metro areas appears to pose contextual and/or environmental risks for cancer, including pediatric leukemia. Future research should explore the possible links between the established environmental risk factors for pediatric leukemia and these nonmetro communities adjacent to metro areas.

Surprisingly, the epidemiological literature on socioeconomic status (SES) and pediatric leukemia is limited. One study that hypothesized that differentials in SES might account for differences in leukemia risk by pediatric age did not find support for that hypothesis [26]. Likewise, a study of the Canadian pediatric population counterintuitively found a slightly lower risk of pediatric leukemia by lower neighborhood per capita income [27]. Additionally, one case-control study on pediatric leukemia found no association with SES [28]. Alternatively, lower SES is a documented risk factor for poorer prognosis and survival among pediatric leukemia patients [13,29]. While not an assessment of risk, the present cross-sectional study observed a significant unadjusted association between U.S. counties with lower median household incomes and pediatric leukemia odds compared to other pediatric cancers. Even so, the multilevel analysis showed that this result was confounded by the significantly greater odds of pediatric leukemia among nonmetro counties adjacent to metro areas, as these counties tend to have lower median household incomes. Therefore, this study’s results do not conflict with prior research that found no differential risk of pediatric leukemia incidence by SES, but this study’s results would also not conflict with a hypothesis of lower neighborhood SES-associated factors being confounded with the relevant environmental risk factors.

A study of pediatric leukemia incidence in the U.S. by race/ethnicity from 1992 to 2013 observed the highest age-adjusted incidence rates for Hispanic whites, and the lowest age-adjusted incidence rates were among non-Hispanic blacks [30]. Asians and whites (both non-Hispanic) had similar incidence rates located between the other two groups. The present study’s case-control design paints a different picture from a different analytic perspective. While adjusting for the other individual and community-level factors, it is non-Hispanic black children who had the greatest odds that a given pediatric cancer would be diagnosed as leukemia, even though the age-adjusted incidence rate was lowest for this group [30]. Non-Hispanic children with other or unknown race likewise had relatively high pediatric leukemia diagnosis odds. Although the age-adjusted incidence rate for Hispanic children was the highest [30], their odds of a pediatric cancer being diagnosed as leukemia were actually intermediate between the high odds groups and the group with the lowest adjusted odds: non-Hispanic white children. The major implication is that a full accounting of racial/ethnic disparities for pediatric cancers should consider not only incidence but also within-group propensities toward certain types of pediatric cancers. As a point of emphasis, although the non-Hispanic black population has had the lowest age-adjusted risk of incident leukemia, this group has also had the greatest adjusted odds that a given pediatric cancer would be diagnosed as leukemia.

The male sex is widely associated with pediatric cancer diagnoses; this association was accentuated among pediatric leukemia patients [31]. The present study similarly found a significantly greater adjusted odds of the male sex among pediatric leukemia patients than among all other pooled pediatric cancers.

Although the risk of a pediatric leukemia diagnosis, especially for acute lymphoblastic leukemia, has been greatest in the one-to-four-year age range [30,32], the adjusted odds that a pediatric cancer diagnosis was leukemia were actually 18 percent lower in the present study for this age range compared to the first year of life. Likewise, even though incident leukemia risk drops off around the age of five years and thereafter, the adjusted odds of pediatric cancer being diagnosed as leukemia were higher in the five-to-14-year age range and were not significantly different from the odds observed among infants. This peculiar result may indicate that although the risk factor exposures for pediatric leukemia may be greatest in the pre-school age range, these exposures may continue into the school age years to a greater extent than the exposures for other pediatric cancers, and the possibly higher in utero exposure due to conceivably greater parental exposure to leukemia risk factors may contribute to the higher relative odds of leukemia among infant cancer diagnoses [7-9].

The external validity of this study for the U.S. pediatric cancer patient population is strong, and there is corroborating evidence that the RUCCs urban/rural population percentages that occur in SEER data are adequate representations of these categories in the U.S. population [18]. The main limitation of this study is that it is a case-control study among pediatric cancer diagnoses and therefore does not inform concerning population-based incidence risk. Nonetheless, the present study has accounted for peer-reviewed pediatric leukemia incidence findings in bringing a fuller context to these results. Also, this potential etiological weakness actually points to a strength of this study: it elucidates the relative, within-group, tendencies of certain individual and community-level characteristics being associated with pediatric leukemia cases. These results reveal how it is possible for a population group with a lower adjusted cancer-specific incidence rate to nonetheless be more consistently impacted by that cancer type than other groups.

## Conclusions

This study found that community-level and patient-level factors both matter simultaneously and independently in setting apart the epidemiological profile of pediatric leukemia from other pediatric cancers. Most notably, counties designated as nonmetro areas adjacent to metro areas were associated with the greatest adjusted odds that a pediatric cancer was diagnosed as leukemia. Future research should explore the extent to which the documented environmental exposure risk factors for pediatric leukemia align geographically with these high-risk areas.

## Acknowledgments

note*: The 21 sources for the SEER 21 dataset included the Alaska Native Tumor Registry, Connecticut, Detroit, Atlanta, Greater Georgia, Rural Georgia, San Francisco-Oakland, San Jose-Monterey, Greater California, Hawaii, Idaho, Iowa, Kentucky, Los Angeles, Louisiana, Massachusetts, New Mexico, New Jersey, New York, Seattle-Puget Sound, and Utah. https://seer.cancer.gov/registries/terms.html

## Data Availability Statement

note†: Access to the SEER 21 data used in this study may be requested at the following website: https://seer.cancer.gov/data/access.html

